# Nicotine and Cannabis Co-Use are Associated with Higher Symptom Burden than Use of Either Drug Alone in Early Psychosis: Results from a Multisite Study

**DOI:** 10.64898/2026.11.03.26367232

**Authors:** Jason Smucny, Ethan M. Campbell, Katherine M. Pierce, Heather B. Ward, Cameron S. Carter, Tara A. Niendam, Tyler A. Lesh

## Abstract

**Background:** Despite their high usage rates and associations with risk for psychosis and earlier age of symptom onset, few studies have examined relationships between nicotine and cannabis co-use and symptoms. Here we examined associations between symptom burden and cannabis/nicotine use/co-use in individuals with early psychosis (EP).

**Methods:** This cross-sectional, multisite cohort study used data from 738 individuals (mean age 22 years, 62% male) from the Early Psychosis Intervention Network (EPI-NET). Relationships between cannabis and/or nicotine use and frequency (over the prior 30 day period) and Modified Colorado Symptom Index (MCSI) scores were examined using mixed effects models, with site as a random factor and age, sex, and cannabis/nicotine use (none, nicotine only, cannabis only, or both) as fixed factors. Exploratory analyses also examined effects of frequency of use (daily vs. less than daily).

**Results:** 59% of the sample used neither nicotine nor cannabis, 17% only nicotine, 9% only cannabis, and 16% both. Significant effects of use were observed on total MCSI score (F = 6.69, *p* < .001) driven by greater symptoms in individuals who used both cannabis and nicotine relative to neither, only nicotine, or only cannabis. More frequent use, furthermore, was associated with higher total score.

**Conclusions:** These results suggest that co-use of cannabis and nicotine is associated with greater symptom burden in EP, which may have implications for treatment strategies. Neuropharmacological mechanisms that may underlie this association are discussed.

## Introduction

Psychotic disorders such as schizophrenia (SZ) are associated with higher prevalence of nicotine and cannabis use relative to unaffected populations. Although estimates vary, rates of cigarette smoking generally range between 60-80% in SZ^1–3^ (∼3x the general population), and approximately one-quarter of people experiencing SZ have a co-occurring cannabis use disorder^4^.

The high rates of cannabis use disorder in psychotic illness are particularly important given evidence that cannabis use increases risk for psychosis. Specifically, lifetime cannabis use is associated with 1.4-fold increase in developing psychosis, and cannabis dependence a 3.4 increased risk ^5^. Cannabis users who develop SZ also have a 2-3 year earlier age of psychosis onset relative to non-users^5^. Using cannabis has also been linked to higher induce positive and negative symptoms of psychosis, such as hallucinations and anhedonia, respectively ^6, 7^. Similarly, smoking tobacco may also increase risk for psychosis, with a relative risk of 2.18 for daily smokers vs. non-smokers as well as a 1.04 years earlier age of psychosis onset in smokers ^8^.

To this point, the great majority of studies examining links between nicotine, cannabis use and psychosis have been conducted by studying relationships with each substance in isolation. However, recent studies suggest that it is important to examine use for both substances simultaneously. Indeed, according to a 2018 study, 48% of people aged 18-25 who smoked tobacco also used cannabis in the last month (compared to 28% of adults > age 25) ^9^; a later (2024) study found that more teens used cannabis and tobacco together than either alone^10^. Interestingly, that study also found that co-users had higher internalizing and externalizing symptoms than non-users and users of just one substance ^10^, suggesting co-use is associated with higher levels of psychopathology.

Amongst work that has examined relationships between nicotine and cannabis co-use and psychosis, a 2023 study by Sanchez-Gutierrez, et al. (2023) ^11^ recruited 1105 patients with first-episode psychosis and 1355 controls, and found that individuals who used tobacco daily as well as cannabis were more likely to have psychosis than non-users or tobacco-only users (odds-ratios = 7.8 for co-users and 3.3 for tobacco only users vs. non-users). Gonzalez-Blanco, et al. (2021) ^12^ found that, in first-episode patients, those who used cannabis and tobacco together had lower age of first prodromal symptoms and higher disorganization symptoms and delusional thinking than those that only used tobacco. Gonzalez-Blanco, et al. (2021) ^12^ did not report relationships between co-users and cannabis only users, however, likely because the sample size for cannabis-only users was low (n = 13). Bello, et al. (2026) ^13^ found that heavy cannabis and light tobacco co-use was associated with the highest risk of conversion to psychosis in individuals at clinical high risk (CHR) for psychosis compared to no use of either substance or either substance alone. Bello, et al. (2026) ^13^ also found that nicotine and cannabis use frequency was associated with symptom severity in CHRs, although they did not find differences in symptom severity when grouping CHRs based on binary classification (e.g., nicotine only, cannabis only, nicotine + cannabis).

These previous studies suggest that use of both cannabis and nicotine together is associated with psychosis, although links to symptom burden are unclear. How nicotine and cannabis co-use relate to symptom severity in individuals with full threshold psychotic illness is particularly poorly understood. Here we leveraged the power of a large, multisite study (the Early Psychosis Intervention Network (EPINET) dataset) to examine these relationships. Based on prior work suggesting nicotine and cannabis co-use is associated with increased risk for psychosis conversion in individuals at CHR ^13^ as well as greater symptom burden in first-episode psychosis ^12^, we hypothesized that use of both substances would be associated with worse overall symptomatology than use of neither or either alone. We also conducted exploratory analyses examining associations with clinical subdomains, frequency of use, alcohol use, and socioeconomic status (SES).

## Methods

### Sample

EPINET (nationalepinet.org) is a network of early psychosis programs across the United States that provide Coordinated Specialty Care (CSC) for psychosis. CSC is a team-based, recovery-oriented approach to care that involves collaborative relationships between the CSC team, individuals seeking care for early psychosis, and their chosen support persons ^14, 15^. CSC programs vary somewhat in their admission criteria, although they typically provide care to individuals aged 12-25 who have experienced psychosis onset within the past 2-5 years ^14, 15^. Notably, all CSC programs participating in EPINET are required to provide core CSC services including therapy, case management, medication management, work and education support, family education, and have an identifiable team lead. All EPINET programs also collect standardized core assessment measures with the goal of collecting data that can be used to inform and improve early psychosis care ^16^.

All available EPINET data was downloaded from the National Data Archive (NDA; nda.nih.gov). Data was gathered from four study repositories: Advancing the Early Psychosis Intervention Network in Texas (EPINET-TX), Connecting First Episode Psychosis Research and Practice through a Learning Health System, Laboratory for Early Psychosis Research (LEAP), and Academic-Community EPINET (AC-EPINET): Mitigating Barriers to Care. The current analysis excluded individuals who used opioids or stimulants in the last 30 days, had missing demographic information (sex/age), had missing symptom information, and individuals diagnosed with substance-induced psychosis or an affective disorder without psychosis. After exclusion criteria were applied, the dataset included information from 47 different sites and 738 participants seeking care at a CSC program nationwide. Only baseline EPINET data were used for analysis.

### Assessment

Symptoms were assessed using the Modified Colorado Symptom Inventory (MCSI) ^17^. The MCSI is a 14 item self-report panel that asks participants how often they experience different symptoms over the past month. Responses are rated as 0 (not at all), 1 (once during the month), 2 (several times during the month), 3 (several times a week), or 4 (at least every day). Symptoms assessed include anxiety, depression, hallucinations, mania, forgetfulness, and suicidal ideations (see Supplementary Material for detailed listing).

Nicotine and cannabis use were assessed with separate yes/no questions (“Have you used cannabis in the past 30 days?”, "Have you used nicotine in the past 30 days?"). Frequency of use in the last 30 days was also assessed (none, less than daily, daily).

As previously for the EPINET dataset ^18^, as a proxy for SES we used health insurance status (private insurance, public insurance, or no insurance). Insurance specified as "other" in the EPINET dataset was considered missing.

### Analysis

Relationships between substance use and average MCSI total score were examined using a mixed effects model with site as a random effect, age and sex as fixed effects, and cannabis/nicotine use (use neither / used nicotine only / used cannabis only / used both) as a fixed effect. Significant main effects of substance use were followed up by post-hoc tests. Significance for post-hoc tests was set to *p* < .05, Sidak-corrected for multiple comparisons. We also examined effect sizes for F tests (η^2^), with .01 < η^2^ < .06 indicating "small", .06 ≤ η^2^ < .14 indicating "medium", and η^2^ ≥ .14 indicating "large" effects (resources.nu.edu/statsresources/eta). To examine potentially confounding effects of SES and antipsychotic use on relationships between nicotine/cannabis use and MCSI total score, SES score (assessed by insurance status; see above) or antipsychotic use (yes/no) were included as fixed factors in exploratory analyses.

As alcohol use was a potential confounder (because individuals who use cannabis/nicotine may be more likely to consume alcohol), a secondary set of exploratory analyses also examined relationships between alcohol and total symptoms in two ways. In the first analysis, nicotine + cannabis non-users were divided into two groups (non-users of any substance and alcohol-only users) prior to being incorporated in the mixed effects model above. Post-hoc tests then compared non-users vs. alcohol-only users. In the second analysis, alcohol use and its interaction with cannabis/nicotine use were included as additional factors in the mixed effects model.

Additional exploratory analyses examined relationships between MCSI "affective", "cognitive", and "positive" symptoms and cannabis/nicotine use were also assessed to determine if associations varied within particular symptom domains. A listing of which questions were included in each of the three domains is shown in Table 1.

**Table 1.** Demographic and clinical information. "Light" cannabis/nicotine use was defined as less than daily (but more than none), and "heavy" use defined as daily use.

| **Measure** | **Mean (SD) or *N* (%)** |
|---|---|
| Total *N* with Complete Data | 738 |
| Age | 21.8 (4.3) |
| Sex |  |
| Male / Female | 454 (61.5) / 284 (38.5) |
| Diagnosis |  |
| SZ | 236 (32.0 |
| SZ-P | 61(8.3) |
| SZ-A | 145 (19.6) |
| PNOS | 133 (18.0) |
| MDD-P | 88 (11.9) |
| BD-P | 75 (10.2) |
| Taking Antipsychotics Y / N / Missing Data | 610 (82.7) / 79 (10.7) / 49 (6.6) |
| Substance Use Last 30 Days Y / N |  |
| Nicotine | 237 (32.2) / 500 (67.8) |
| Cannabis | 182 (24.7) / 555 (75.3) |
| Alcohol | 169 (22.9) / 569 (77.1) |
| Substance Use Combinations Last 30 Days |  |
| No Alcohol, Cannabis, Or Nicotine Use | 388 (52.6) |
| Only Alcohol | 46 (6.2) |
| Only Nicotine | 122 (16.5) |
| Only Cannabis | 67 (9.1) |
| Both Cannabis and Nicotine | 115 (15.6) |
| Nicotine/Cannabis Use Frequency Last 30 Days |  |
| None | 434 (58.8) |
| Light Nicotine Only | 33 (4.5) |
| Light Cannabis Only | 46 (6.2) |
| Heavy Nicotine Only | 84 (11.4) |
| Heavy Cannabis Only | 21 (2.8) |
| Light Nicotine and Light Cannabis | 26 (3.5) |
| Heavy Nicotine and Light Cannabis | 40 (5.4) |
| Light Nicotine and Heavy Cannabis | 6 (.8) |
| Heavy Nicotine and Heavy Cannabis | 36 (4.9) |
| Missing Data | 12 (1.6) |
| Nicotine/Cannabis Use Frequency (Combined) |  |
| None | 434 (58.8) |
| Use One Lightly | 79 (10.7) |
| Use One Heavily | 105 (14.2) |
| Use Both, At Least One Less Than Heavily | 72 (9.8) |
| Use Both Heavily | 36 (4.9) |
| Missing Data | 12 (1.6) |
| Health Insurance Status |  |
| Private Insurance | 258 (35.0) |
| Public Insurance | 300 (40.7) |
| No Insurance | 110 (14.9) |
| Other or Missing Data | 70 (9.5) |
| Modified Colorado Symptom Inventory (MSCI) Total Score (n = 738) | 20.9 (13.9) |
| 1. How Often Nervous? | 2.2 (1.4) |
| 2. How Often Depressed? | 1.8 (1.4) |
| 3. How Often Lonely? | 1.8 (1.5) |
| 4. How Often Others Said You Were Paranoid? | 1.0 (1.3) |
| 5. How Often Hear Voices? | 1.4 (1.6) |
| 6. How Often Indecisive? | 2.0 (1.5) |
| 7. How Often Concentration Problems? | 2.0 (1.5) |
| 8. How Often Behavior Felt Strange? | 1.6 (1.5) |
| 9. How Often Felt Out of Place? | 1.7 (1.6) |
| 10. How Often Forgetful? | 1.6 (1.4) |
| 11. How Often Racing Thoughts? | 1.6 (1.5) |
| 12. How Often Suspicious/Paranoid? | 1.4 (1.4) |
| 13. How Often Felt Like Hurting Yourself/Suicidal? | .5 (.9) |
| 14. How Often Felt Like Hurting Someone Else? | .3 (.8) |
| Avg Affective Symptoms (#s 1, 2, 3, 9, 11, 13, 14) | 1.4 (1.0) |
| Avg Cognitive Symptoms (#s 6, 7, 10) | 1.9 (1.3) |
| Avg Positive Symptoms (#s 4, 5, 8, 12) | 1.3 (1.2) |

Finally, exploratory analyses were also performed analyzing relationships between frequency of use and symptoms. The sample was divided into 5 groups for this analysis (used none, used nicotine (only) or cannabis (only) less than daily, used nicotine (only) or cannabis (only) daily, used both cannabis and nicotine but did not use both daily, used both cannabis and nicotine daily). Relationships with frequency of use were conducted using mixed effects models with site as a random effect and sex/age/frequency of use as fixed effects. Effect sizes were also examined using the guidelines above. Significant main effects of frequency of use were followed up by a planned linear contrast (use none < use one less than daily < use one daily < use both but did not use both daily < use both daily). Significance for the linear contrast was set to *p* < .05. Relationships between use/co-use for nicotine and cannabis and frequency of use for each drug were examined using chi-square tests, with significance also set to *p* < .05.

## Results

### Demographics

Demographic and clinical information as well as cannabis and nicotine use data are presented in Table 1. After applying exclusion criteria and removing individuals with missing substance use and/or clinical information, 738 individuals across 47 sites were included in analysis. 505 individuals did not have MCSI data and were thus excluded from analysis. Comparisons between substance use patterns for individuals with vs. without MCSI scores are presented in Supplementary Table 1. No significant differences were observed in nicotine/cannabis use between those participants with and without symptom data.

MCSI frequency scores were between one and two (i.e., between once during the month and several times during the month) for most questions and for each symptom domain (affective, cognitive, and positive (see Methods)). 59% of individuals did not use either cannabis or nicotine in the last 30 days, 17% used nicotine only, 9% used cannabis only, and 16% used both cannabis and nicotine.

### Symptom Profiles based on Cannabis/Nicotine Use

Total MSCI scores depending on cannabis and/or nicotine use are illustrated in Figure 1 (covariate-adjusted parameter estimates extracted from ANOVA models) and Supplementary Figure 1 (raw means); statistical comparisons are presented in Tables 2a and 2b. Briefly, a significant effect of use was observed on MCSI total symptom score, although the effect size was small (η^2^ = .030). The effect was driven by significantly higher symptoms in individuals that used cannabis and nicotine together vs. those that used neither and those that used nicotine only. Individuals that used only cannabis did not differ symptomatically from any other substance use profile.

**Figure 1.**
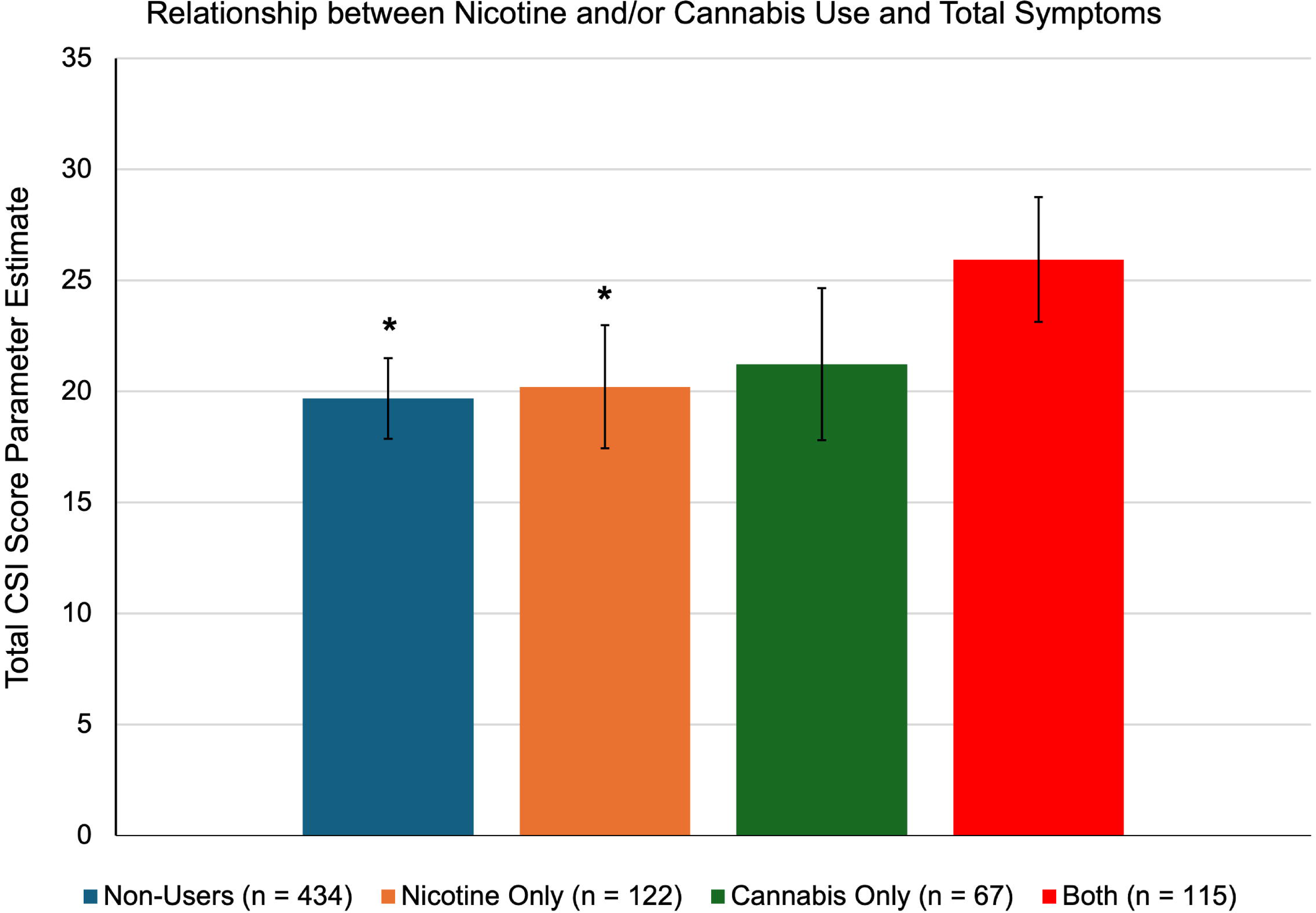
Association between nicotine and cannabis use and Colorado Symptom Inventory (MCSI) total score. Parameter estimates are adjusted for age, sex, and site. \**p* < .05 vs. individuals who use both.

**Table 2.** Overall ANOVA F test (top) and post-hoc test results (bottom) for comparisons of Modified Colorado Symptom Index (MSCI) score between individuals who use neither nicotine nor cannabis, nicotine only, cannabis only, or both. *Significant difference (*p* < .05, Sidak-corrected). MSE = mean square error.

| **Overall Model** | **F** | **df** | **MSE** | **Partial η<sup>2</sup>** | ***p*** |
|---|---|---|---|---|---|
| MCSI Total Symptoms (Sum) | 6.69 | 3, 686 | 163.18 | .028 | < .001 |
| **Post-Hoc Comparison** | **Estimated Mean Difference** | **SE** | **95% CI** | ***p*<sub>Sidak</sub>** |  |
| Non-Use vs. Nicotine Only | -.04 | .10 | -.30 – .23 | >.99 |  |
| Non-Use vs. Cannabis Only | -.11 | .13 | -.44 – .22 | .94 |  |
| Non-Use vs. Use Both | -.45 | .10 | -.71 – -.18 | < .001* |  |
| Nicotine Only vs. Use Both | -.41 | .12 | -.73 – -.09 | .005* |  |
| Cannabis Only vs. Use Both | -.34 | .15 | -.73 – .05 | .13 |  |
| Nicotine Only vs. Cannabis Only | -.07 | .15 | -.46 – .31 | >.99 |  |

Exploratory analyses of symptom domains are illustrated in Supplementary Figures 2 (parameter estimates) and 3 (raw means); statistical comparisons are presented in Supplementary Tables 2a and 2b. Similar between-group results were observed for affective, cognitive, and positive symptoms, although the only significant between-group difference for positive symptoms was observed between non-users and individuals who used both.

### Cannabis/Nicotine + Alcohol Use

When individuals who used neither nicotine nor cannabis were segregated into groups that also did not use alcohol or used only alcohol, the effect of substance use on total MCSI symptom score remained significant (F_4, 685_ = 5.04, mean square error (MSE) = 163.40, *p* < .001, η^2^ = .029). Amongst participants that used neither nicotine nor cannabis, post-hoc tests found no difference between those that did not vs. those that did use alcohol (did not use alcohol estimate = 19.61, used alcohol estimate = 20.31, mean difference = -.70, standard error (SE) = 2.13, *p* > .99).

When alcohol use was included an additional factor in mixed models, no effect of alcohol use was observed (F_1,685_ = 1.51, MSE = 163.62, *p* = .22, η^2^ = .002). The interaction effect between nicotine/cannabis use and alcohol use was also non-significant (F_3_, _682_ = .33, MSE = 163.62, p = .81, η^2^ = .001).

### Symptom Profiles based on Cannabis/Nicotine Use Frequency

MSCI total scores based on cannabis and/or nicotine use frequency are illustrated in Figure 2 (covariate-adjusted estimates) and Supplementary Figure 4 (raw means). A significant main effect of cannabis/nicotine use frequency and MCSI total symptoms was observed (F_4, 673_ = 6.19, MSE = 161.53, *p* < .001, partial η^2^ = .04). The planned linear contrast of frequency of use relationships (use none < use nicotine-only or cannabis-only less than daily < use nicotine-only or cannabis-only daily < use both but did not use both daily < use both daily) was also significant (linear contrast estimate = 6.45, 95% CI = 3.26 – 9.65, *p* < .001).

**Figure 2.**
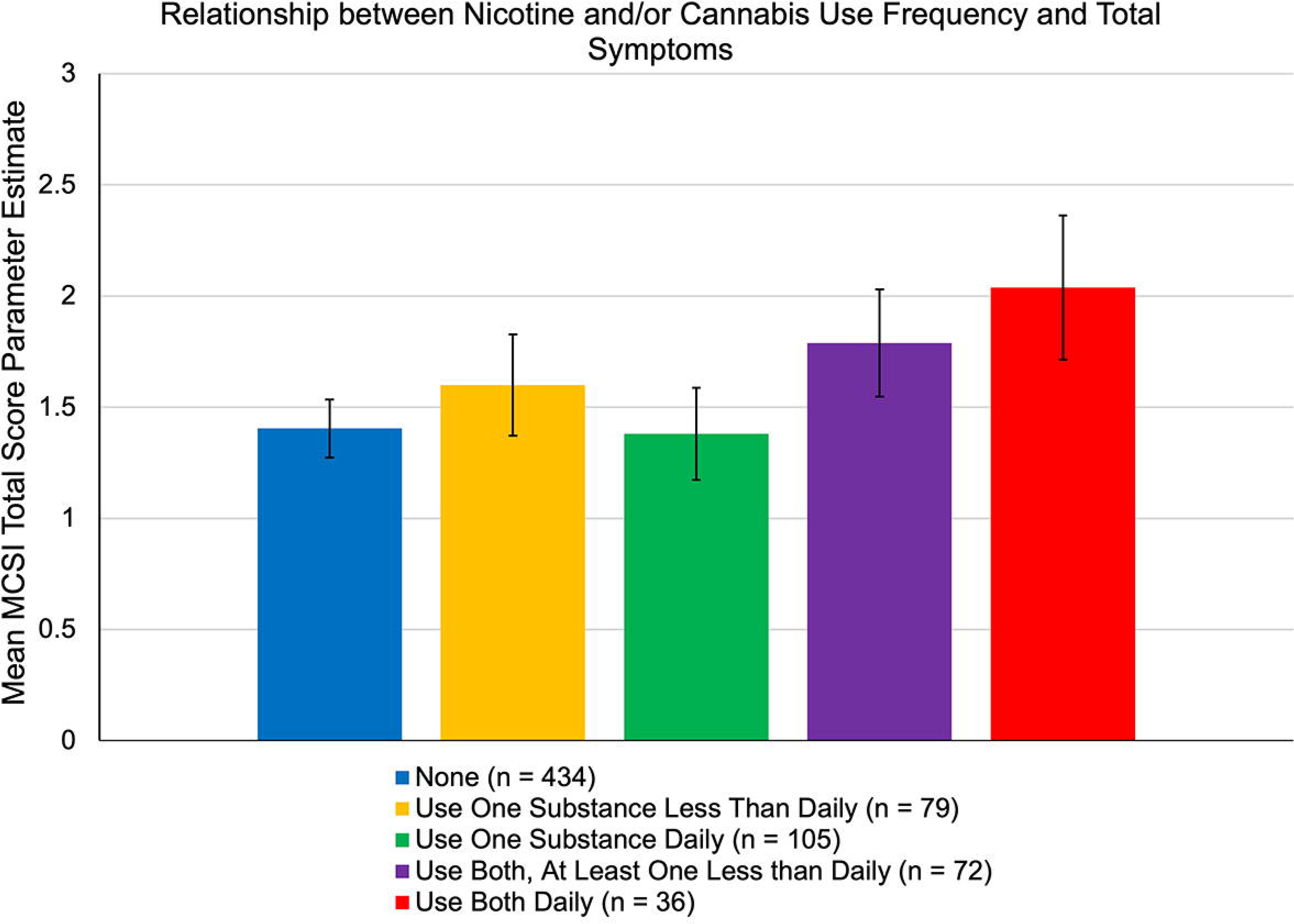
Association between frequency of nicotine and cannabis use and Colorado Symptom Inventory (MCSI) total score (top left) and MCSI symptom domains (top right, bottom left, and bottom right). Parameter estimates are adjusted for age, sex, and site.

Results from exploratory analyses of MSCI symptom domains based on use frequency are presented are presented in Supplementary Table 3. Parameter estimates and raw means are presented in Supplementary Figures 5 and 6, respectively. Briefly, significant main effects of cannabis/nicotine use frequency were observed for all symptom domains. Planned linear contrasts of frequency of use relationships were also significant for all symptom domains.

### Frequency of Use for Individuals with Co-Use vs. Single Substance Use

Frequency of use data was missing for 12 participants. Amongst the 175 individuals using nicotine who also had frequency of use data available, no difference was found between nicotine only users vs. individuals who used both nicotine and cannabis in nicotine use frequency (χ^2^ = .022, *p* = .88). Specifically, amongst 117 individuals who used nicotine only, 33 (28.2%) used the drug lightly and 84 (71.8%) used the drug heavily (i.e., daily). Amongst 110 individuals who used both nicotine and cannabis, 32 (29.1%) used the drug lightly and 78 (70.9%) used the drug heavily.

Similarly, amongst the 179 individuals using cannabis and who also had frequency of use data available, no difference was observed between those who used cannabis only and those who used both nicotine and cannabis in cannabis use frequency (χ^2^ = .91, *p* = .34). Specifically, amongst the 67 individuals who used cannabis only, 46 (68.7%) used the drug lightly and 21 (31.3%) used the drug heavily. Amongst the 112 individuals who used both nicotine and cannabis, 69 (61.6%) used the drug lightly and 43 (38.4%) used the drug heavily.

### Socioeconomic Status Effects

A question regarding medication affordability was used as a proxy for SES (data presented in Table 1). A plurality of individuals had public insurance (41%). When SES was added as a factor to a model predicting MCSI total score based on nicotine/cannabis use, the SES effect was non-significant (F_2, 615_) = 1.06, MSE = 163.30, *p* = .35, η^2^ = .00). The effect of nicotine/cannabis usage also remained significant (F_2, 615_ = 6.96, MSE = 163.30, *p* < .001, η^2^ = .033).

### Antipsychotic Effects

Amongst individuals with available mediation status information, 89% were taking antipsychotics (Table 1). No significant effects of antipsychotic use status were observed on models based on nicotine/cannabis use.

## Discussion

Overall, the results of this study suggest that, after controlling for effects of age and sex, nicotine and cannabis co-use is associated with greater overall symptom severity in EP compared to use of neither substance or either substance alone. This difference was not dependent on the symptom domain examined (affective, cognitive, or positive). Symptom scores were not different between individuals who did not use any substances versus those who used only alcohol. Alcohol use was also not significantly associated with symptoms and showed no interactions with nicotine/cannabis use on symptoms. Controlling for SES effects also did not affect the relationship between nicotine/cannabis use and MCSI total score. Linear contrast of dose relationships also suggested that, after controlling for effects of age and sex, those individuals that used both substances daily had the highest symptom burden. The lack of significant relationships between use/co-use and frequency of use for either drug, however, suggests the differences in symptom severity between users and co-users is unlikely to be due to differences in frequency of use between users and co-users.

While prior work has shown that nicotine and cannabis co-use is associated with greater odds of psychosis^11^ or conversion from CHR ^13^ relative to isolated use of either drug, these results show that the combined drug effect may continue to impact clinical outcomes by exacerbating symptom burden in individuals who experience threshold psychosis. This is consistent with research showing greater symptom severity among nicotine and cannabis co-use relative to nicotine without cannabis use ^12^, but also leverages a larger sample of those who use cannabis but not nicotine to suggest that co-use is more deleterious than single use alone.

Combined with results from prior literature, the results also suggest that EP may be a period of particularly high vulnerability to symptom exacerbation related to nicotine and cannabis co-use over the course of illness (compared to those with chronic illness). Specifically, in CHR individuals, Bello et al. (2026) ^13^ found higher rates of conversion in in participants who used both substances vs. those that used none or either alone, as well as relationships between frequency of use for both nicotine and cannabis and symptom severity. In contrast, in a review of studies examining mostly chronic (average age ∼40 years) people with schizophrenia, Sabe et al. (2020)^19^ found that positive symptoms were only marginally elevated in nicotine and cannabis users vs. those that used neither drug. The subgroup analyses from this review, however, were limited as all cannabis users also used cannabis, precluding comparisons vs. nicotine or cannabis only users.

While directionality cannot be inferred between co-use and symptom severity in EP from the present study, their co-use may have synergistic effects that may ultimately exacerbate symptoms of psychosis. A large body of preclinical work indicates that the nicotinic cholinergic and endocannabinoid systems exhibit functional interdependence, where pharmacological or genetic alteration of one system produces neurochemical and behavioral changes in the other ^20^. Both cannabis and nicotine elicit acute increases in dopamine release ^21, 22^, and co-use may synergistically sensitize dopamine firing ^23^. Given that hyperdopaminergia is plausibly linked to psychosis symptoms, co-use of these drugs may feasibly lead to emergence and exacerbation of these symptoms ^24^. In addition, while the expression of cannabinoid-1 receptors on cortical glutamatergic and GABAergic neurons ^25^ suggest excitatory-inhibitory balance plays a role in cannabis-associated psychosis ^26^, these neurons also express nicotinic acetylcholine receptor subtypes ^27^. The co-use of nicotine and cannabis may thus contribute to even more pronounced excitatory-inhibitory dysregulation. Additionally, Rabin et al. (2025) ^28^ found that cannabis and tobacco co-use were associated with elevations in fatty acid amide hydrolase compared to cannabis use only, suggesting increased metabolism of the endocannabinoid anandamide associated with nicotine exposure. The downregulation of anandamide is another possible mechanism for the present results, as it is thought to serve a neuroprotective role and inversely correlates with cross-domain symptom severity in EP ^29^. Structurally, co-use in adolescent and young adult populations has been tied to aberrations in cortical white matter tracts ^30, 31^, changes that could underly morphological contributions to symptom severity ^32^. Finally, it should be noted that the preponderance of the evidence (including genomic studies) suggest that cannabis use is a causal mechanism of psychosis ^33^, although it is unclear if nicotine (when combined with cannabis) synergizes this relationship.

In regard to our examination of SES effects, our results suggest SES did not affect relationships between use and symptoms, although only a SES proxy (health insurance status) was available to assess SES across the majority of individuals in the dataset. The impact of socioenvironmental factors like early childhood adversity have been implicated in the relationship between cannabis use and psychosis^34, 35^, and their inclusion could explain whether co-use is involved in a multi-hit pathway^36^ to symptom exacerbation. Additional research is thus needed to more completely understand the relationships between environmental adversity (e.g., low SES), nicotine/cannabis use, and symptom severity in psychotic illness.

While strengthened by the use of a large multisite cohort of EP participants to resolve differences between groups, the present study had several limitations. Firstly, antipsychotic dose information was not available, which could affect the results due to pharmacokinetic drug interactions^37, 38^ and differential neurometabolic responses as a function of cannabis use^39^. Additionally, while past 30-day use frequency was used to delineate levels of use, a more comprehensive substance use inventory would be beneficial for future research. For cannabis use, the age of initiation^40^, accelerating vs. decelerating use patterns^41^, quantity/potency^42^, and cumulative exposure^43^ are implicated in clinical outcomes. Cannabis strain information may have also been useful, as some evidence suggests that the ratio of tetrahydrocannabinols (exacerbating ^44^) to cannabidiol (protective ^45^) may affect how cannabis use influences psychosis symptoms. A retrospective, more detailed account of substance use history could better explain how co-use develops relative to illness onset and contributes to symptom burden.

In conclusion, the results of this study suggest that co-use of nicotine and cannabis is associated with greater psychosis symptom burden in EP relative to use of neither substance of either alone. The study also suggests a dose effect, where daily use of both substances was associated with the most severe symptoms. These findings expand on prior work suggesting that co-use may be a synergistic risk factor for psychosis in CHR by demonstrating symptom burden after illness onset in EP. The prior literature and present study emphasize that co-use should be specifically accounted for as a specific risk factor in populations at-risk for or diagnosed with psychosis. One possible avenue for future research is to use longitudinal approaches (including causal models) to examine temporal relationships between use and symptoms. Future work can also test specific mechanistic hypotheses based on prior work on co-use in other populations such as bioassays for endocannabinoids like anandamide and stress hormones^46^, imaging methods that probe dopamine and glutamate/GABA, cannabinoid receptors and behavioral mechanisms such as reward responsivity and neurocognitive performance.

## Data Availability

All data produced are available online from the National Data Archive (nda.nih.gov).

http://nda.nih.gov

## Acknowledgements

The authors thank the study site coordinators and providers of EPINET for allowing for the use of their data repository on nda.nih.gov, as well as the associated patients and their families. Author J.S. is supported by grant K-01 MH125096 from the National Institutes of Mental Health.

