## Supplementary Material for "Nicotine and Cannabis Co-Use are Associated with Higher Symptom Burden than Use of Either Drug Alone in Early Psychosis: Results from a Multisite Study"

**Supplementary Methods**

*Colorado Symptom Inventory*

The Colorado Symptom Inventory (CSI) is a 14-item self-report measure that asks participants about the frequency they have experience symptoms over the past month (never, once, several times, several times per week, at least every day). The 14 CSI questions are:

1. How often have you felt nervous, tense, worried, frustrated, or afraid?
2. How often have you felt depressed?
3. How often have you felt lonely?
4. How often have others told you that you acted "paranoid" or "suspicious"?
5. How often did you hear voices, or hear and see things that other people didn’t think were there?
6. How often did you have trouble making up your mind about something, like deciding where you wanted to go or what you were going to do, or how to solve a problem?
7. How often did you have trouble thinking straight or concentrating on something you needed to do (like worrying so much or thinking about problems so much that you can’t remember or focus on other things)?
8. How often did you feel that your behavior or actions were strange or different from that of other people?
9. How often did you feel out of place or like you did not fit in?
10. How often did you forget important things?
11. How often did you have problems with thinking too fast (thoughts racing)?
12. How often did you feel suspicious or paranoid?
13. How often did you feel like hurting or killing yourself?
14. How often have you felt like seriously hurting someone else?

**Supplementary Table 1**. Comparison of substance use patterns for individuals with vs. without symptom data. "Light" cannabis/nicotine use was defined as less than daily (but more than none), and "heavy" use defined as daily use.

| **Measure** | **Mean (SD) or *N* (%) with Symptom Data** | **Mean (SD) or *N* (%) without Symptom Data** | **t or χ^2^ (*p*)** |
| --- | --- | --- | --- |
| *N* | 738 | 505 | – |
| Substance Use Last 30 Days Y/N | | | |
| Nicotine | 237 (32.2) / 500 (67.8) | 155 (30.7) / 350 (69.3) | .28 (.60) |
| Cannabis | 182 (24.7) / 555 (75.3) | 133 (26.3) / 372 (73.7) | .45 (.51) |
| Alcohol | 169 (22.9) / 569 (77.1) | 113 (22.6) / 388 (77.4) | .02 (.89) |
| Substance Use Combinations Last 30 Days |  | | 2.74 (.60) |
| No Alcohol, Cannabis, Or Nicotine Use | 388 (52.6) | 261 (51.7) |  |
| Only Alcohol | 46 (6.2) | 29 (5.7) |  |
| Only Nicotine | 122 (16.5) | 82 (16.2) |  |
| Only Cannabis | 67 (9.1) | 60 (11.9) |  |
| Both Cannabis and Nicotine | 115 (15.6) | 73 (14.5) |  |
| Nicotine/Cannabis Use Frequency |  | | 6.59 (.58) |
| None | 434 (58.8) | 290 (57.4) |  |
| Light Nicotine Only | 33 (4.5) | 27 (5.3) |  |
| Light Cannabis Only | 46 (6.2) | 32 (6.3) |  |
| Heavy Nicotine Only | 84 (11.4) | 48 (9.5) |  |
| Heavy Cannabis Only | 21 (2.8) | 21 (4.2) |  |
| Light Nicotine and Light Cannabis | 26 (3.5) | 12 (2.4) |  |
| Heavy Nicotine and Light Cannabis | 40 (5.4) | 22 (4.4) |  |
| Light Nicotine and Heavy Cannabis | 6 (.8) | 8 (1.6) |  |
| Heavy Both | 36 (4.9) | 26 (5.1) |  |
| Missing Data | 12 (1.6) | 19 (3.8) |  |
| Nicotine/Cannabis Use Frequency (Combined) |  | | 1.01 (.91) |
| None | 434 (58.8) | 290 (57.4) |  |
| Use One Lightly | 79 (10.7) | 59 (11.7) |  |
| Use One Heavily | 105 (14.2) | 69 (13.7) |  |
| Use Both, At Least One Less Than Heavily | 72 (9.8) | 42 (8.3) |  |
| Use Both Heavily | 36 (4.9) | 26 (5.1) |  |
| Missing Data | 12 (1.6) | 19 (3.8) |  |

**Supplementary Table 2**. Overall ANOVA F tests (top) and post-hoc test results (bottom) for comparisons of Modified Colorado Symptom Index (MSCI) symptom domain scores between individuals who use neither nicotine nor cannabis, nicotine only, cannabis only, or both. *Significant difference (*p* < .05, Sidak-corrected). MSE = mean square error.

| **Comparison** | **F** | **df** | **MSE** | **Partial η^2^** | ***p*** |
| --- | --- | --- | --- | --- | --- |
| Affective Symptoms (Mean) | 7.18 | 3, 686 | .82 | .030 | <.001 |
| Cognitive Symptoms (Mean) | 4.95 | 3, 686 | 1.37 | .021 | .002 |
| Positive Symptoms (Mean) | 3.80 | 3, 686 | 1.20 | .016 | .010 |

| **Post-Hoc Comparison** | **Estimated Mean Difference** | **SE** | **95% CI** | ***p*_Sidak_** |
| --- | --- | --- | --- | --- |
| *Affective Symptoms* | | | | |
| Non-Use vs. Nicotine Only | -.06 | .10 | -.32 – .20 | .99 |
| Non-Use vs. Cannabis Only | -.13 | .12 | -.46 – .20 | .89 |
| Non-Use vs. Use Both | -.46 | .10 | -.73 – -.20 | < .001* |
| Nicotine Only vs. Use Both | -.40 | .12 | -.72 – -.08 | .006* |
| Cannabis Only vs. Use Both | -.33 | .15 | -.72 – .05 | .13 |
| Nicotine Only vs. Cannabis Only | -.07 | .14 | -.45 – .31 | > .99 |
| *Cognitive Symptoms* | | | | |
| Non-Use vs. Nicotine Only | -.02 | .13 | -.36 – .31 | > .99 |
| Non-Use vs. Cannabis Only | -.22 | .16 | -.64 – .21 | .69 |
| Non-Use vs. Use Both | -.48 | .13 | -.82 – -.13 | .002* |
| Nicotine Only vs. Use Both | -.46 | .16 | -.87 – -.04 | .023* |
| Cannabis Only vs. Use Both | -.26 | .19 | -.76 – .24 | .67 |
| Nicotine Only vs. Cannabis Only | -.19 | .19 | -.69 – .30 | .88 |
| *Positive Symptoms* | | | | |
| Non-Use vs. Nicotine Only | -.01 | .12 | -.33 – .31 | > .99 |
| Non-Use vs. Cannabis Only | -.00 | .15 | -.40 – .40 | > .99 |
| Non-Use vs. Use Both | -.40 | .12 | -.72 – -.08 | .007* |
| Nicotine Only vs. Use Both | -.39 | .15 | -.77 – .00 | .052 |
| Cannabis Only vs. Use Both | -.40 | .18 | -.86 – .07 | .14 |
| Nicotine Only vs. Cannabis Only | .01 | .17 | -.45 – .47 | > .99 |

**Supplementary Table 3**. Top: Overall ANOVA F tests and for MSCI symptom domains comparisons between individuals based on frequency of nicotine and/or cannabis use (including users only). Bottom: Results for the linear contrast examining relationships between MCSI symptom domains and frequency of use (use none < use one less than daily < use one daily < use both but did not use both daily < use both daily). MCSI = Modified Colorado Symptom Inventory, MSE = mean square error.

| **MCSI Score** | **F** | **df** | **MSE** | **Partial η^2^** | ***p*** |
| --- | --- | --- | --- | --- | --- |
| Affective Symptoms (Mean) | 6.61 | 4, 673 | .81 | .04 | < .001 |
| Cognitive Symptoms (Mean) | 4.91 | 4, 673 | 1.36 | .03 | < .001 |
| Positive Symptoms (Mean) | 3.48 | 4, 673 | 1.20 | .02 | .008 |

| **MCSI Score** | **Linear Contrast Estimate** | **95% CI** | ***p*** |
| --- | --- | --- | --- |
| Affective Symptoms (Mean) | .51 | .28 – .73 | < .001 |
| Cognitive Symptoms (Mean) | .43 | .14 – .72 | .004 |
| Positive Symptoms (Mean) | .41 | .13 – .68 | .004 |

**Supplementary Figure 1.** Association between nicotine and cannabis use and Modified Colorado Symptom Inventory (MCSI) total raw score.

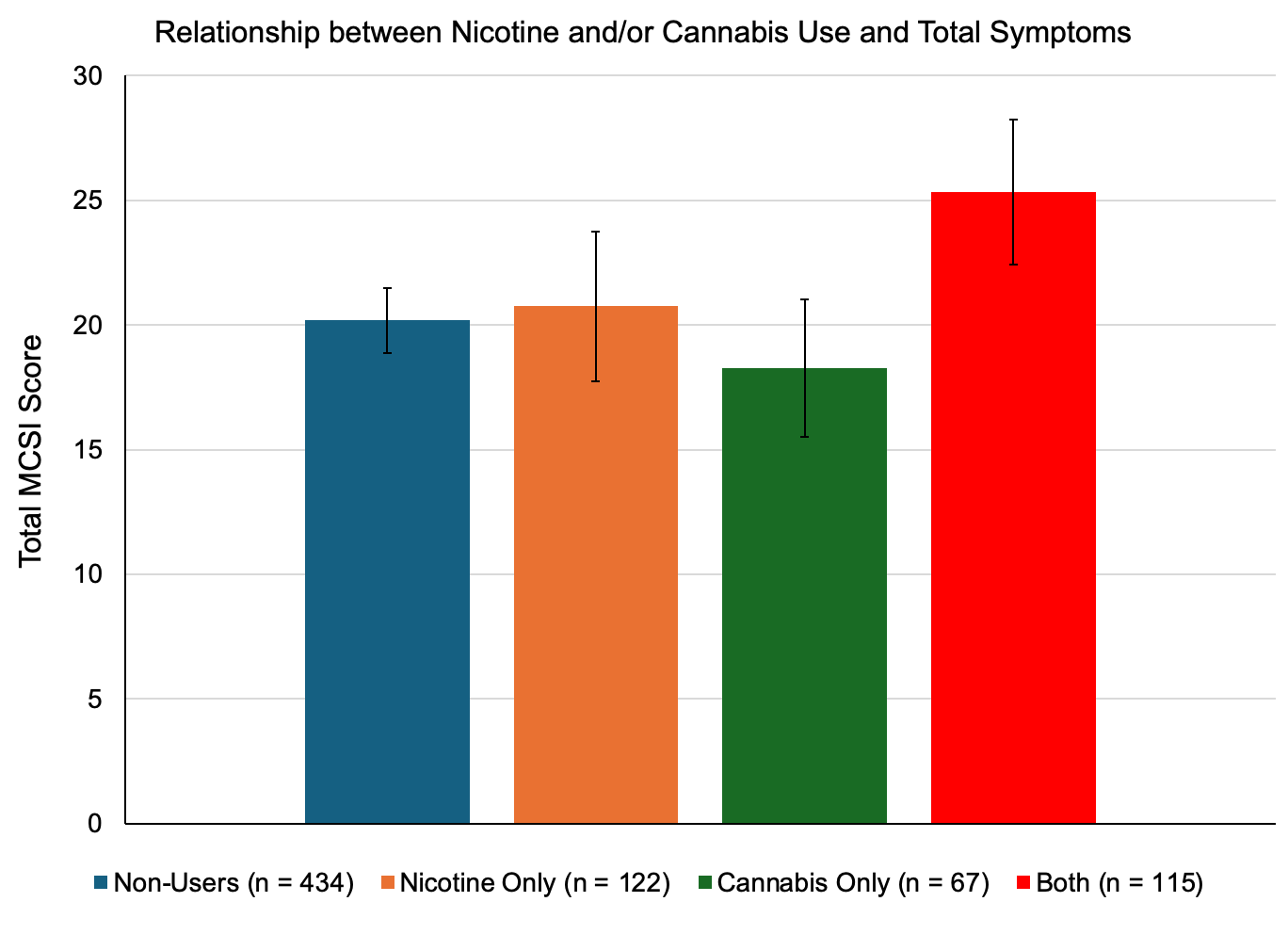

**Supplementary Figure 2.** Association between nicotine and cannabis use and Modified Colorado Symptom Inventory (MCSI) symptom domain model parameter estimates. Parameter estimates were adjusted for age, sex, and site. **p* < .05 vs. individuals who use both.

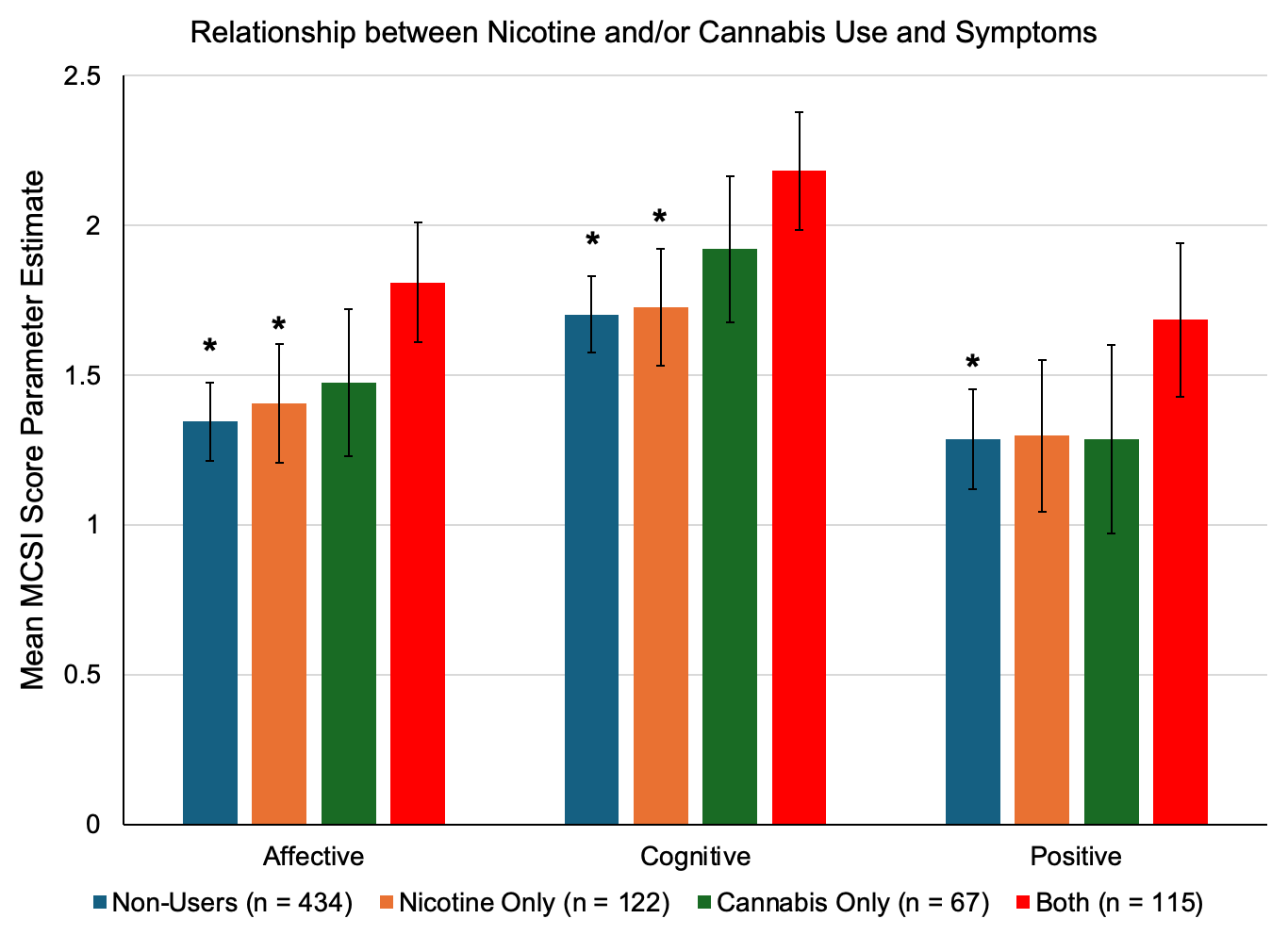

**Supplementary Figure 3.** Association between nicotine and cannabis use and Modified Colorado Symptom Inventory (MCSI) symptom domain raw scores.

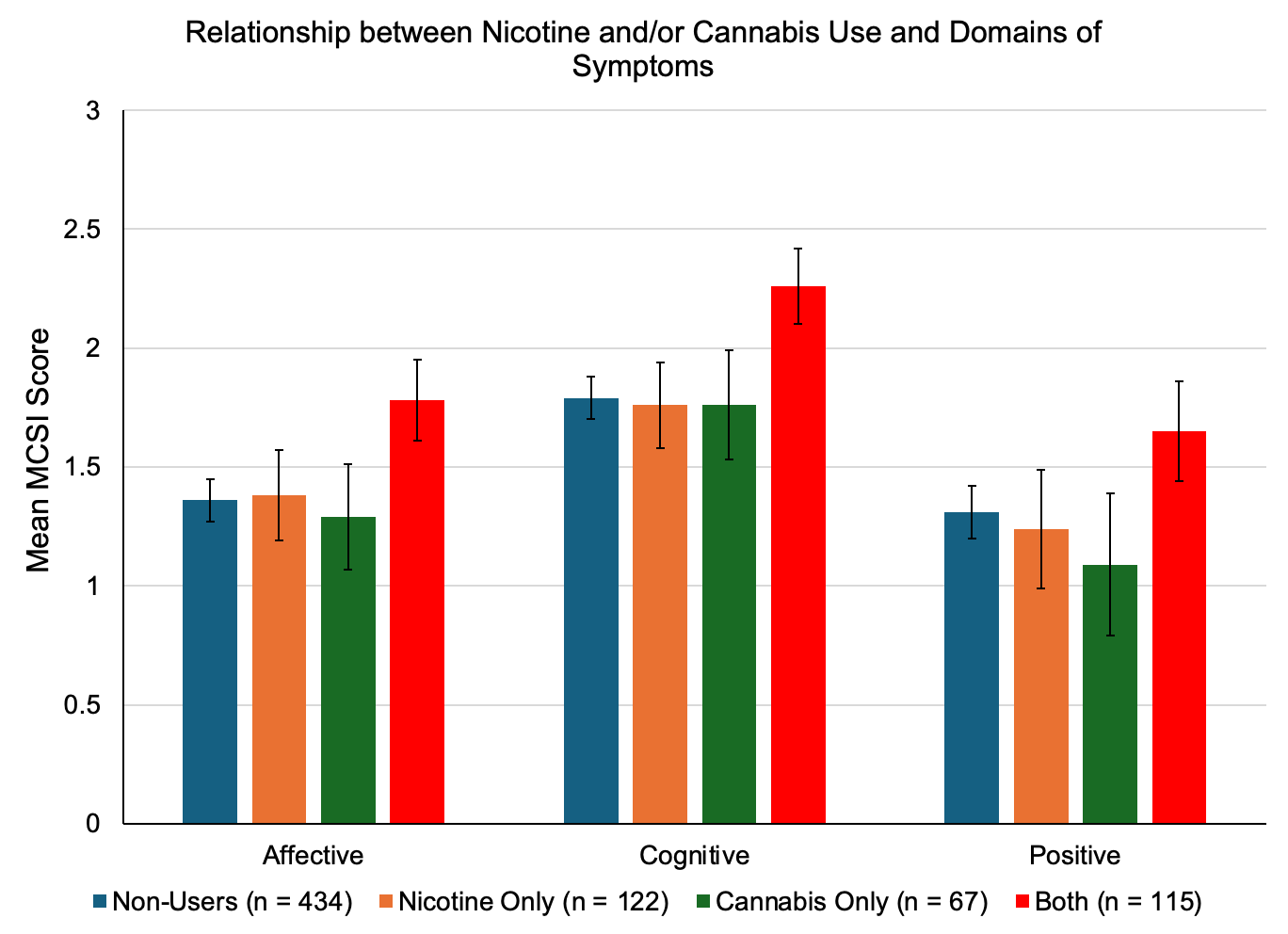

**Supplementary Figure 4.** Association between frequency of nicotine and cannabis use and Modified Colorado Symptom Inventory (MCSI) total raw score.

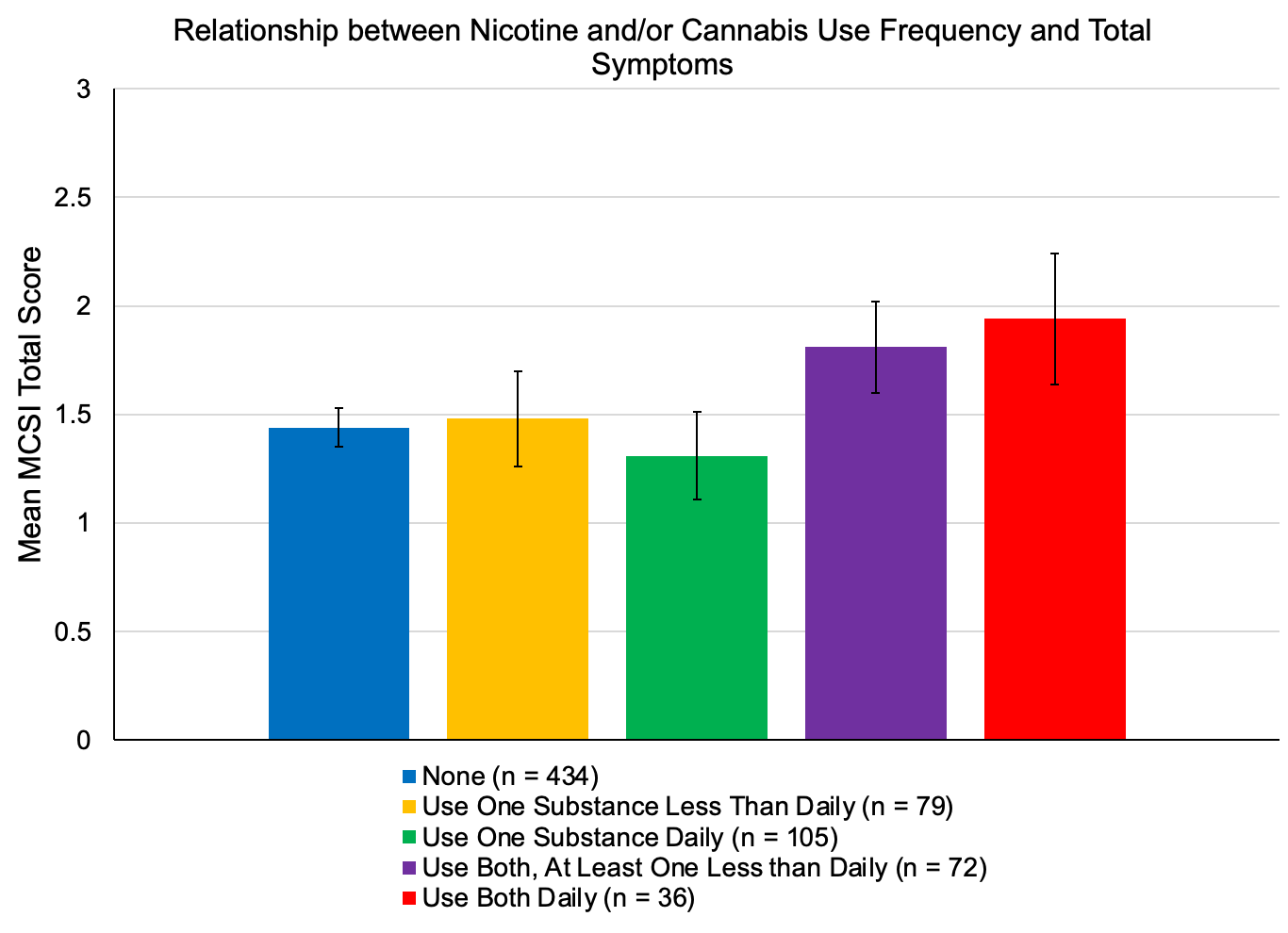

**Supplementary Figure 5.** Association between frequency of nicotine and cannabis use and Modified Colorado Symptom Inventory (MCSI) symptom domain mean domain model parameter estimates. Parameter estimates were adjusted for age, sex, and site.

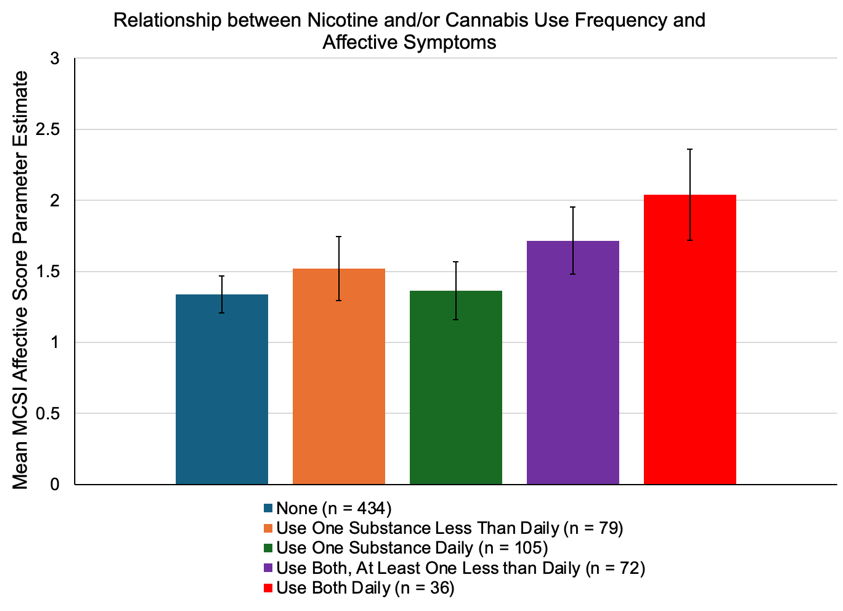

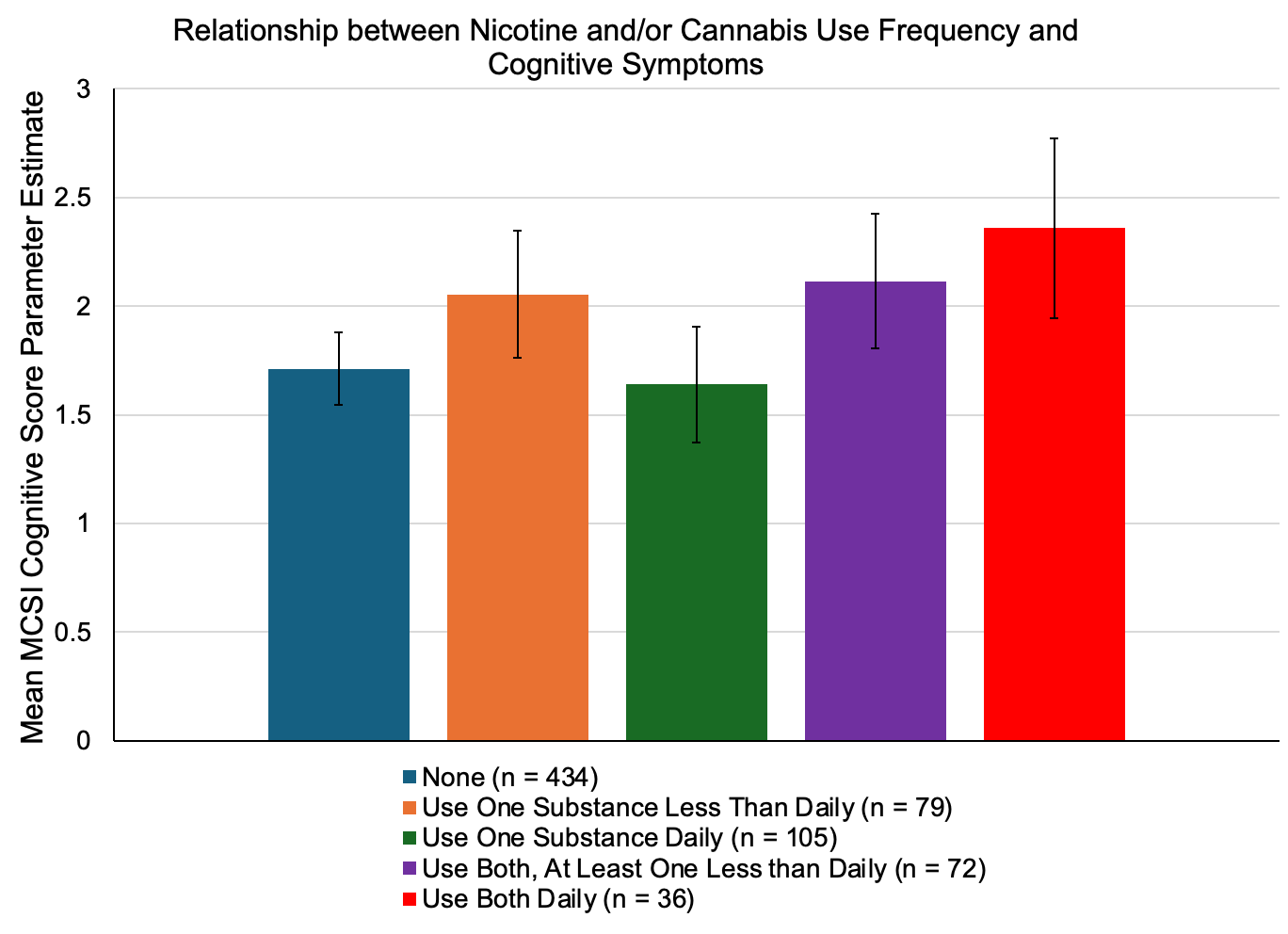

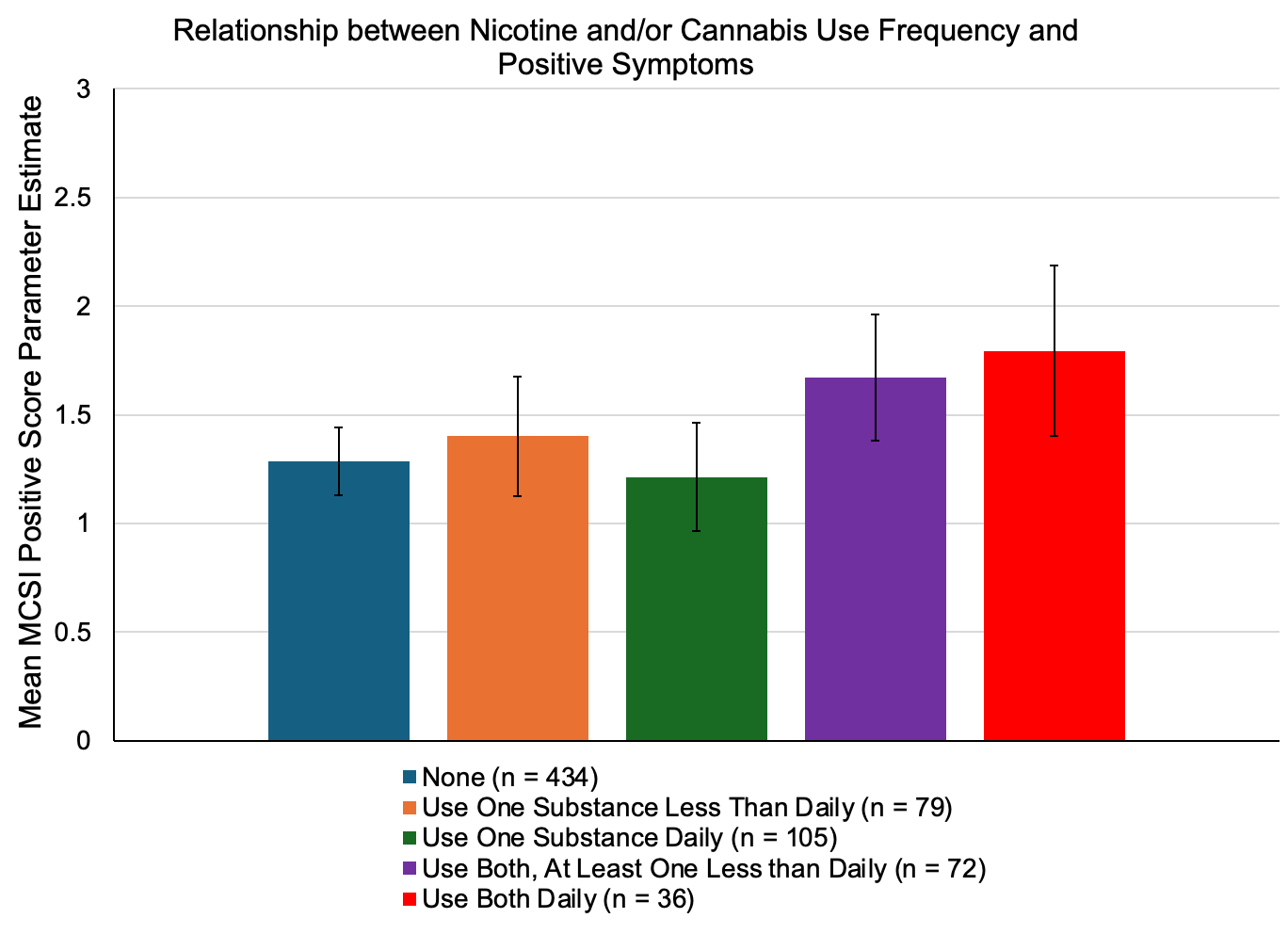

**Supplementary Figure 6.** Association between frequency of nicotine and cannabis use and Modified Colorado Symptom Inventory (MCSI) symptom domain mean raw scores.

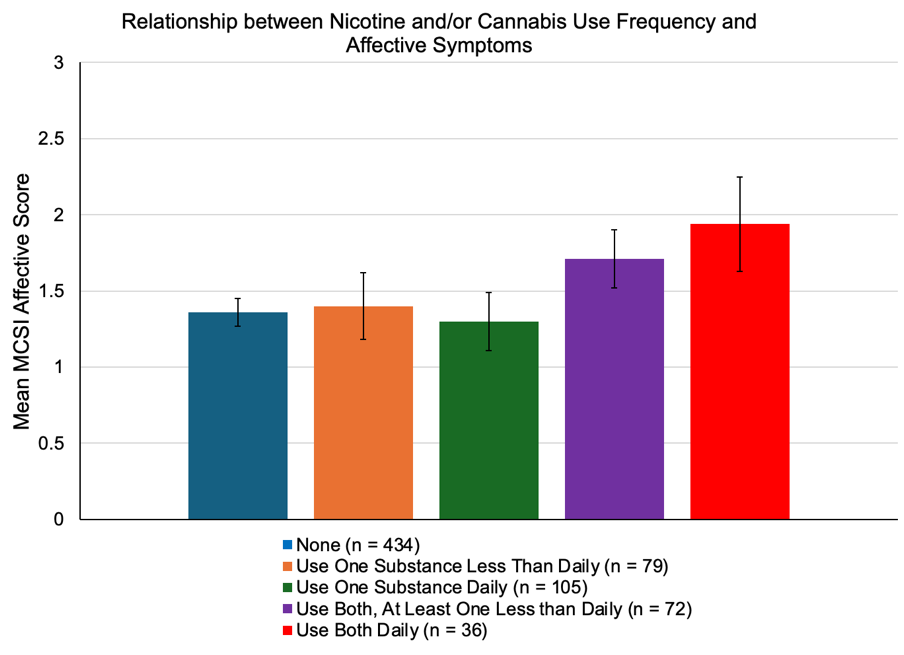

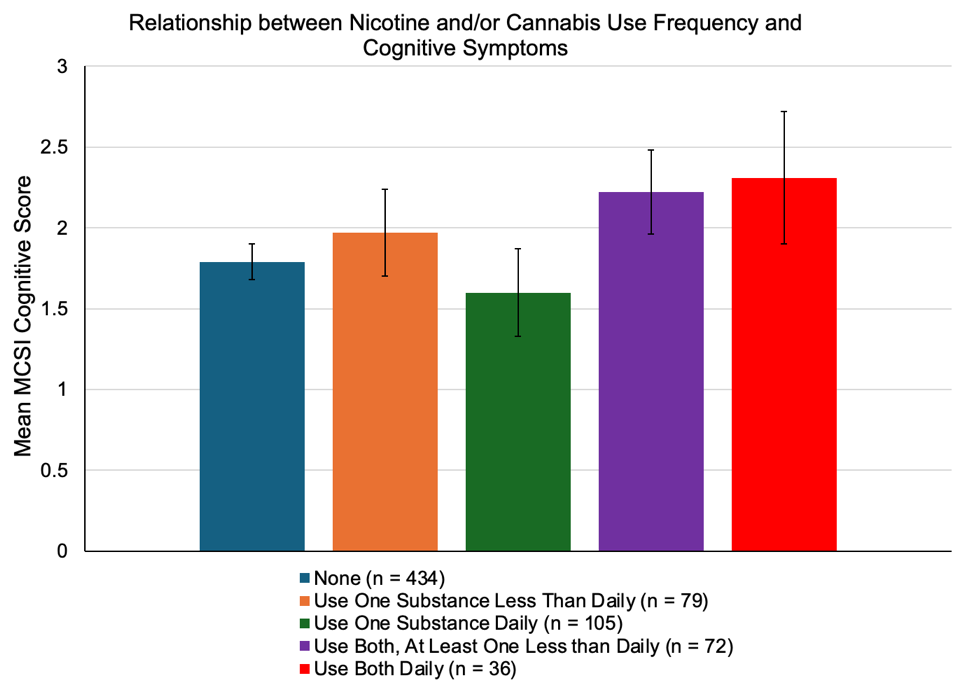

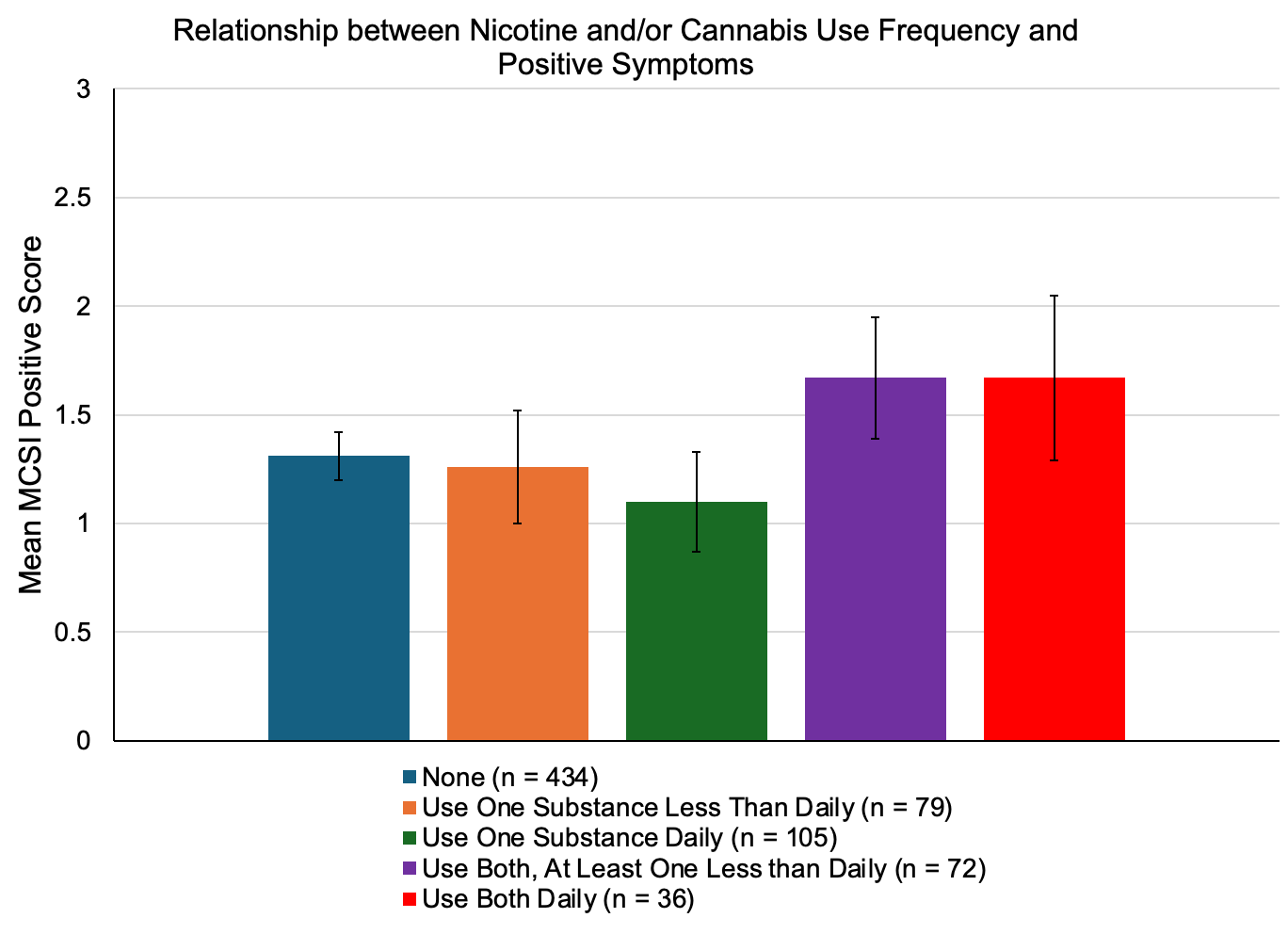
